# Shared Medical Appointments in Heart Failure For Post-hospitalization Follow-up: A Randomized Controlled Trial

**DOI:** 10.1101/2024.03.04.24303754

**Authors:** Tracey H. Taveira, Lisa B. Cohen, Sherry K. Laforest, Karen Oliver, Melanie Parent, Rene Hearns, Sherry L. Ball, Sandesh Dev, Wen-Chih Wu

## Abstract

**Background:** Shared medical appointments (SMAs) in heart failure (HF) are medical visits where several patients with HF meet with multidisciplinary providers at the same time for efficient and comprehensive care. It is unknown whether HF-SMAs can improve overall and cardiac health status for high-risk patients discharged with HF.

**Methods:** A 3-site, open-label, randomized-controlled-trial was conducted. Participants within 12 weeks of HF hospitalization were randomized to receive either HF-SMA or usual HF clinical care (usual-care) on a 1:1 ratio. The HF-SMA team, which consisted of a nurse, nutritionist, psychologist, nurse practitioner and/or a clinical pharmacist, provided four 2-hour session HF-SMAs that met every other week for 8 weeks. Primary outcomes were the overall health status measured by EQ5D-VAS and cardiac health status by KCCQ, 180 days post-randomization.

**Results:** Of the 242 patients enrolled (HF-SMA n=117, usual-care n=125, mean age 69.3±9.4 years, 71.5% white, 94.6% male), 84% of participants completed the study (n=8 HF-SMA and n=9 usual-care patients died). After 180 days, both HF-SMA and usual-care participants had similar and significant improvements from baseline in KCCQ, but only HF-SMA participants had significant improvements in EQ5D-VAS (mean change = 7.2 ± 15.8 in HF-SMA versus -0.4 ± 19.0 points in usual-care, p<0.001).

**Conclusion:** Both HF-SMA and usual care in HF participants achieved significant improvements in cardiac health status, but only a team approach through HF-SMA achieved significant improvements in overall health status. A larger population and a longer follow-up time are needed in future studies to evaluate re-hospitalization and death outcomes.

**Clinical Perspective:**

- This randomized controlled trial represents the first multi-site study to rigorously assess the outcomes of a multidisciplinary team intervention in a group medical clinic setting to improve patient-centered self-reported outcomes of high-risk patients with a recent heart failure hospitalization.
- Shared medical appointments can improve cardiac specific and overall health status in high-risk patients with heart failure.

## Introduction

Heart failure (HF) poses a significant burden on healthcare systems, necessitating a comprehensive and patient-centered multidisciplinary team approach to its management.^1,2^ The conventional one-provider to one-patient medical appointment model often falls short in delivering holistic care due to time constraints for patients and providers in one visit, limited time and resources of patients to attend multiple appointments for providers of different disciplines and expertise, and difficulty in care coordination amongst the providers of different disciplines.^3,4^ Shared medical appointments (SMAs) are medical visits in which several patients with a common complex disease have clinic at the same time with a team of providers of different disciplines, to provide timely, comprehensive, and efficient care.^5^ SMAs in HF are a unique opportunity in which providers of different disciplines, such as nutrition, social work, psychology, and a medication prescriber with HF expertise, can meet with HF patients in one setting, address distinct care barriers, and shift the burden of care coordination and communication from the patient to the provider team, while fostering peer support and self-management education.^6^ However, randomized-controlled trial evidence supporting the impact of SMAs in HF on patient-centered outcomes is limited, especially for complex HF patients with recent hospitalizations. Therefore, our objective was to conduct a randomized-controlled trial to compare HF-SMA versus usual HF clinical care in patient-centered outcomes, such as overall and cardiac-specific health status, and HF self-care, in patients with a recent HF hospitalization. We hypothesized that patients discharged with HF randomized to a HF-SMA will experience better overall health status, as measured by the European Quality of Life Visual Analog Scale (EQ5D-VAS), cardiac health status, measured by Kansas City Cardiomyopathy Questionnaire (KCCQ), and heart failure self-care, compared to usual care, after 180 days.^7–9^

## Methods

### Trial design and oversight

We conducted a 3-site open label randomized-controlled efficacy trial (RCT) of parallel design to evaluate the effect of our HF-SMA group visit model (intervention arm, n=117) versus usual care (control arm, n=125) for participants within 12 weeks of a HF hospitalization. The study was conducted at three urban VA Medical Centers in Providence, RI (n=92), Phoenix, AZ (n=19) and Cleveland, OH (n=131). Study recruitment began in September 2015 and concluded in June 2019. Details of the trial design have been previously published.^10^ This trial was registered with Clinicaltrials.gov public database, a registry and results database of publicly and privately supported clinical studies of human participants (ClinicalTrials.gov Identifier: NCT02481921). The Institutional Review Board and Research and Development Committees at the Providence, Phoenix, and Cleveland Veterans Affairs Medical Centers approved the protocol. All study procedures were conducted in accordance with the ethical standards of the Helsinki Declaration of 1975. Written informed consent was obtained from each participant prior to randomization. All authors had access to and participated in the interpretation of the analyzed data. The first, second and senior authors prepared the initial draft of the manuscript, which was reviewed and edited by all authors. All the authors vouch for the completeness and accuracy of the data; and vouch for the fidelity of the trial to the protocol. This trial was sponsored by a Veterans Affairs Health Service Research and Development Merit Review Grant IIR 14-293.

### Participants

Patients were eligible if they there were 18 years of age or older, whom, in the previous 12 weeks, were discharged from the hospital with a primary diagnosis of HF or received intravenous diuretics in the emergency room or other ambulatory care settings for HF and were able to provide informed consent. Participants were excluded if they were: 1) unable to attend group sessions due to either psychiatric instability (e.g., acutely suicidal, psychotic) or organic brain injury (e.g. severe dementia, encephalopathy) that precluded self-reporting on health status; 2) discharged to hospice or nursing home facilities for long term care, or patients with a code status of comfort-measures-only; 3) recipients of heart transplant or ventricular assist devices; 4) receiving intravenous inotropic infusions, 5) pregnant, or 6) with end-stage renal disease on dialysis or end-stage liver disease; since the goals of care and treatment management strategies would be very different compared to general HF patients after acute care. Of note, participants were not excluded if they were currently or previously participated in HF education classes, support groups or HF individual provider clinics as these co-interventions are often present in an optimal “HF care” setting.

### Data Collection

In person study visits and electronic health record reviews were conducted to obtain baseline demographics and medical co-morbidities in the Charlson co-morbidity index^11^, laboratory testing and to gather information regarding emergency room and hospitalization utilization. In addition to in-person visits, monthly telephone calls and chart reviews were conducted to obtain information regarding participation in a HF co-intervention (i.e. cardiology, HF one to one clinic and HF education classes), emergency room use, hospitalizations or death. Medical records for reported hospitalizations, emergency room visits or deaths were obtained for event confirmation.

### Randomization

Participants were randomized in blocks of 4 on a 1:1 ratio using a computerized random sequence generator by a call-in method to the central coordinating site (Providence VA) and stratified based on the following variables; 1) current co-intervention enrollment in a HF clinic, or HF education program versus none; 2) ≤2 versus >2 hospitalization in the last 6 months; 3) left ventricular ejection fraction <40% versus ≥ 40% and; 4) study site.

### HF-SMA Intervention

The HF-SMA teams at each site consisted of a nurse, nutritionist, psychologist, nurse practitioner and/or a clinical pharmacist prescriber and included four 2-hour sessions that met every other week for 8 weeks. The first-half of each session (30 minutes) consisted of HF education based on curriculum outlined by the HF Society of America (https://hfsa.org/patient-hub/heart-failure-basics-patients-guide). The second half consisted of behavioral strategies for HF self-management and monitoring (15 min) followed by an individualized cardiovascular focused physical assessment and pharmacologic interventions in the group setting (60-90 min). All SMAs included a provider with prescriptive capability; they were not merely didactic classes. Usual Care Arm

All participants received standard HF care as dictated by their cardiologists, HF specialists and primary care providers. All participants were encouraged to schedule an appointment with their physicians/health care providers within 14 days of a HF discharge. HF clinic was present in all three enrollment sites as part of the usual HF care.

### Outcomes

Outcome data were collected by patient self-report at baseline, 90-days, and 180-days to minimize missing data due to drop-outs. The co-primary outcome was the change from baseline, at 180 days post-randomization, in the current overall health status, as measured by the self-rated EQ5D-VAS which ranges from 0-100 with 0 being “the worst health you can imagine” and 100 being “the best health you can imagine”.^7^ The second co-primary outcome was the change in cardiac health status as measured by the KCCQ.^8,9^ The KCCQ is a 23-item instrument validated in stable and decompensated HF patients with preserved and reduced EF.^8^ The questionnaire reflects HF-specific health status over the prior two weeks. The domains of KCCQ are physical limitation, symptoms, self-efficacy, social limitation, and quality of life. A minimal clinically important score change for both the EQ5D-VAS and KCCQ surveys is 5 points, with a 10- and 20-point change reflecting moderate and large clinical changes, respectively.^12^

Secondary outcomes of interest included HF self-care behavior as measured by the HF Self-Care of Heart Failure Index (SCHFI). ^13,14^ The SCHFI is a 22-item self-administered instrument composed of 3 scales: self-care maintenance, management, and confidence. The survey assesses diet, exercise, medication use, when to call the health provider, keeping doctor’s appointments, weight monitoring, and recognition of change in health status or symptoms. Each scale is scored separately and has a range of 0-100. A score of ≥70 on each scale is considered adequate self-care, though benefit occurs at even lower levels. A score change of half standard deviation (∼8 points) is considered a clinically relevant change.^13,14^ The management subscale of the SCFHI focuses specifically on a patient’s competence in managing their HF on a day-to-day basis. This subscale includes items related to medication adherence, symptom recognition and response to symptoms. The self-care maintenance subscale is an important component of the SCHFI because it provides insight into a patient’s ability to engage in self-care activities, adhere to their prescribed self-care regimen and prevent worsening of heart failure symptoms. Aspects covered in the self-care maintenance subscale include medication and dietary adherence, fluid management, engagement in regular physical activity, follow-up with providers, ability to recognize and monitor heart failure symptoms and lifestyle modifications such as tobacco cessation. The confidence subscale assesses the level of self-efficacy that individuals with HF have in performing self-care activities.

Hospitalizations, emergency room utilizations and death were tracked during monthly phone calls and chart reviews for all study participants.

#### Sample size and power consideration

Using a conservative dropout rate of 15% over the 180 days of follow-up and assuming a baseline mean EQ5D-VAS and KCCQ scores of 60, with a common standard deviation of 13 for the study arms and a 2-sided significance level of 0.05, we had >80% power to detect a clinically significant change in EQ5D-VAS and KCCQ of 5 points with an effective sample size of 108 participants per group (15% less the total sample).^15^ A 2-sided p-value of < 0.05 was considered significant.

### Statistical methods

All analyses were done on an intention-to-treat basis and performed using STATA/SE version 11.2 software (StataCorp LP, College Station, TX). Baseline characteristics were compared between the HF-SMA and usual care utilizing t-tests for continuous variables and chi-square tests for discrete variables. Data were reported as means ± standard deviation when continuous and as percentages when in categories.

#### Primary Outcomes

For the primary analyses, we used generalized estimating equation modeling which accounts for repeated measures within patients over time to compare the change in overall and cardiac health status from baseline between patients in the HF SMA and usual HF care groups, adjusting for randomization stratifiers (co-intervention enrollment in a HF clinic, HF education program versus none; ≤2 versus >2 hospitalization in the last 6 months; left ventricular ejection fraction <40% versus ≥ 40% and; study site) and for imbalanced baseline characteristics (hypertension).^16^ The within group change from baseline to 180 days of follow-up was compared using paired t-testing. We used all available participant data; no observations were deleted because of missing follow-up data. Complete survey data for both the EQ5D-VAS and KCCQ were available for 83.9% and 83.8% for HF-SMA participants at 90 and 180 days respectively. Complete survey data for both the EQ5D-VAS and KCCQ were available for 88.0% at 90 days usual care participants. At 180 days 84% of usual care participants completed the EQ5D-VAS and 84.8% completed the KCCQ surveys.

#### Secondary Outcomes

Similar to above, generalized estimating equation modeling was used to compare the change from baseline over the 180 days of follow-up in the 3 SCHIFI scales: self-care maintenance, management, and confidence between patients in the HF SMA and usual HF care groups. Paired t-tests were used to compare the within group change from baseline to 180 days of follow-up for each study arm.

In order to understand potential heterogeneity of the treatment effects, linear regression with multiplicative interaction between the randomization stratification variables (current co-intervention enrollment in a HF clinic or HF education program versus none; ≤2 versus >2 hospitalization in the last 6 months; left ventricular ejection fraction <40% versus ≥ 40% and; study site) and the randomization arm was used to assess for effect modification, if any, on the change in KCCQ and EQ5D-VAS scores.

Cox proportional hazards modeling was used to compare the time to death or hospitalization over the 180 days of follow-up between the two treatment arms. We tested the proportionality of hazards assumption for the Kaplan-Meier curve by analysis of the Schoenfeld Residuals.^17^ The proportional hazards assumption was confirmed for the time to rehospitalization or death analysis (p=0.85).

## Results

Of the 1219 patients who were screened for eligibility and were contacted for enrollment, 242 (16.5%) provided informed consent and were randomly assigned to one of the study arms. Of the 242 patients randomized, 83.8% (n=98) in the HF-SMA and 84.8% (n=106) in the usual HF care arms completed the study and were analyzed for the primary outcome (Figure 1).

**Figure 1:**
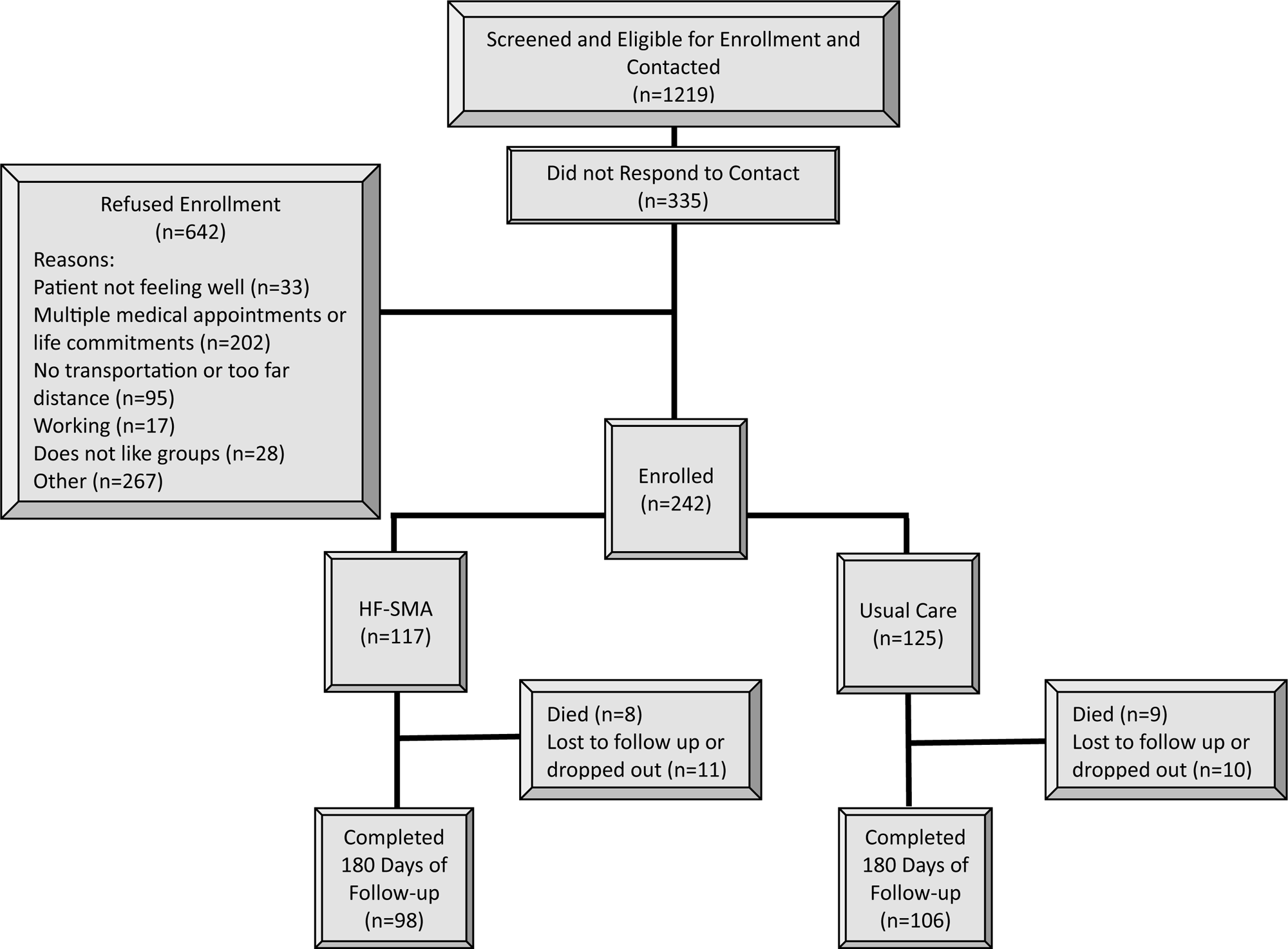
Study Flow Diagram

The characteristics of the study population are shown in Table 1. The study cohort had a mean age of 69.3 ± 9.4 years. Overall, 36.4% of participants had a baseline left ventricular ejection fraction of <40%. Baseline demographic and comorbid conditions were well balanced between the study arms with the exception of hypertension (89.7% for HF-SMA versus 96.8% usual HF care, p=0.03). The mean baseline EQ5D-VAS (64.6 ± 20.1 in HF-SMA versus 66.8 ± 22.4 in usual care, p=0.42) and KCCQ (60.4 ± 23.9 in HF-SMA versus 58.7 ± 24.3 in usual care, p=0.59) scores were also similar. A similar number of participants were receiving care in the individual provider HF clinic (17.1% in HF-SMA versus 13.6% in usual care, p=0.45).

**Table 1.**
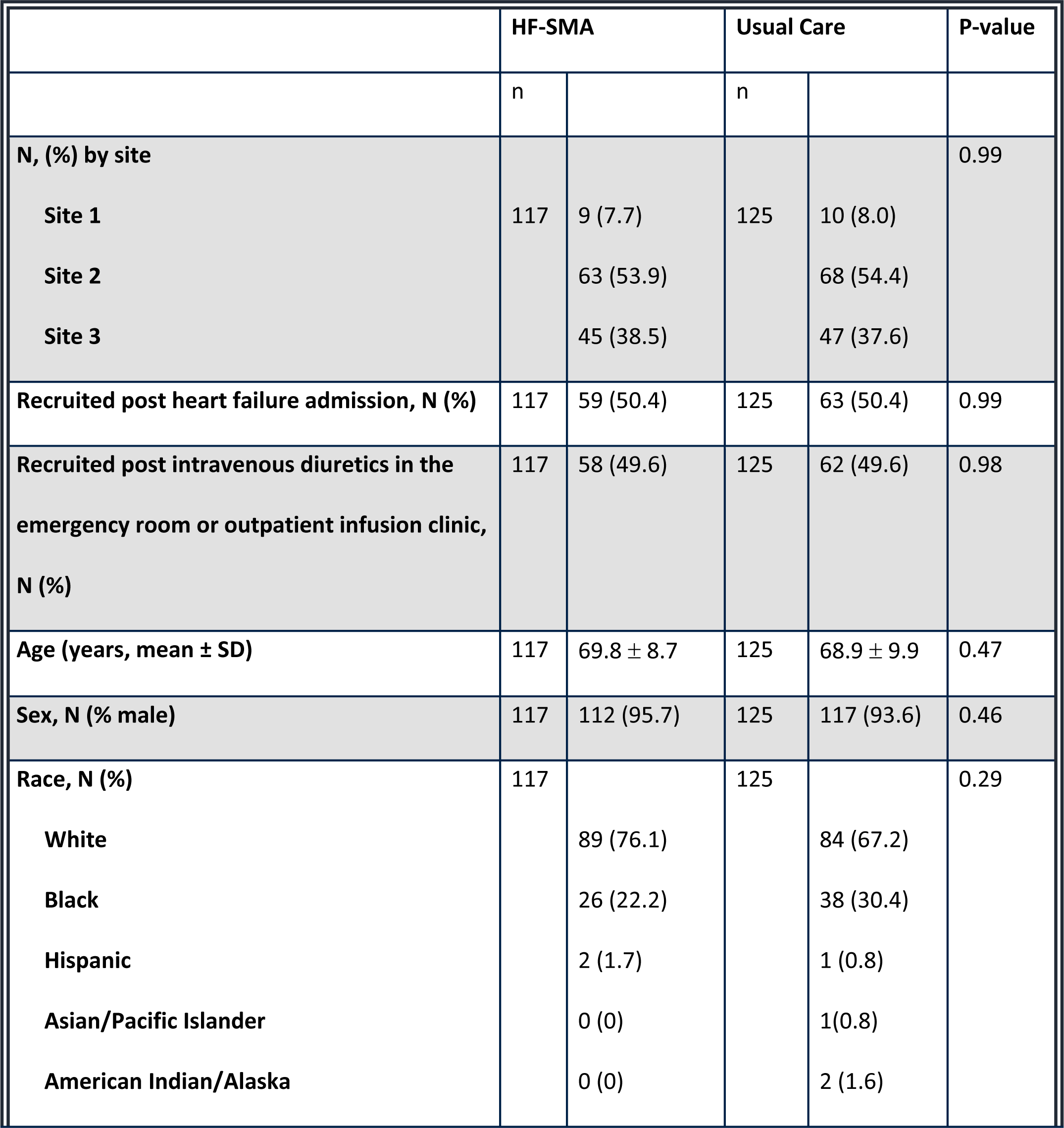
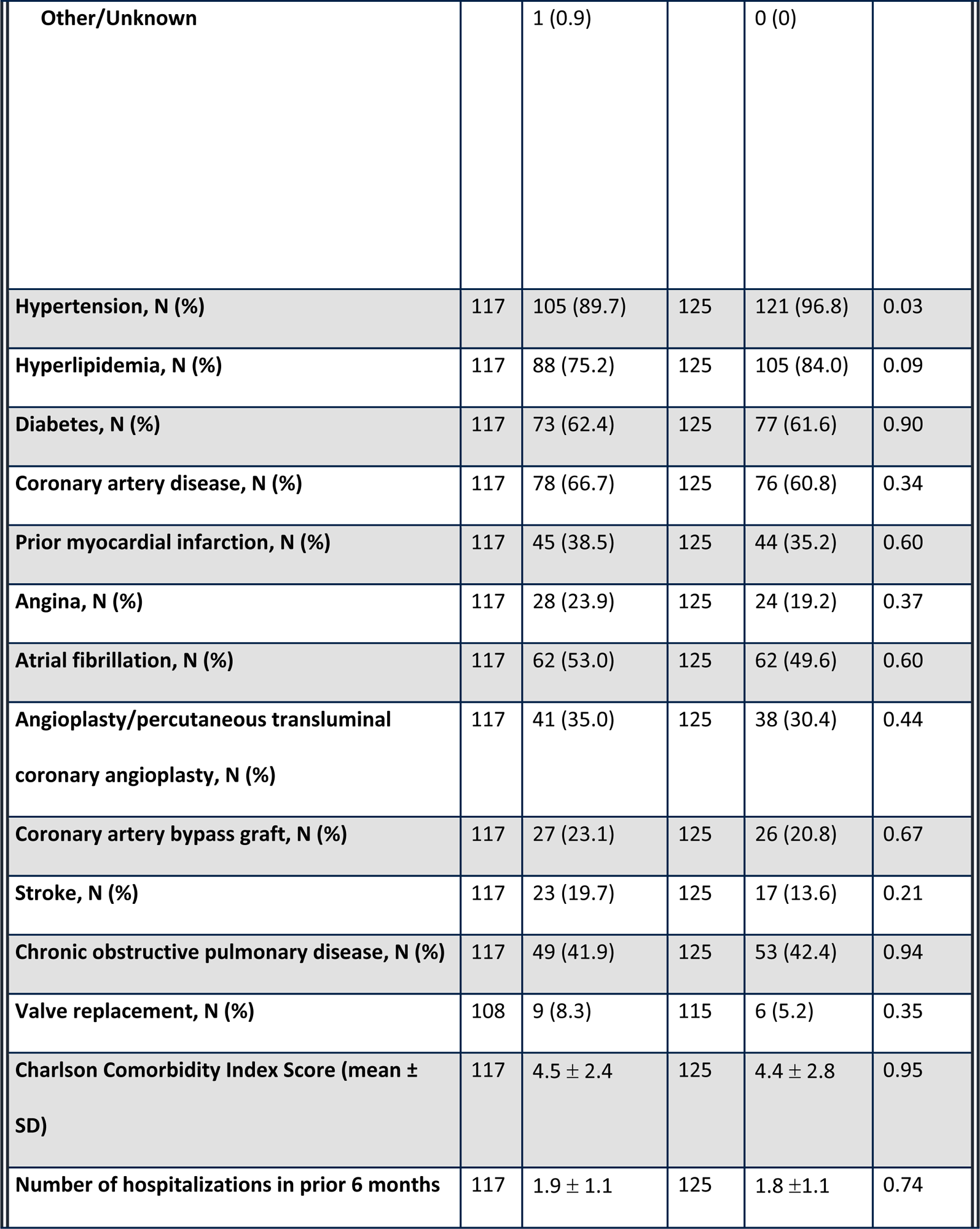
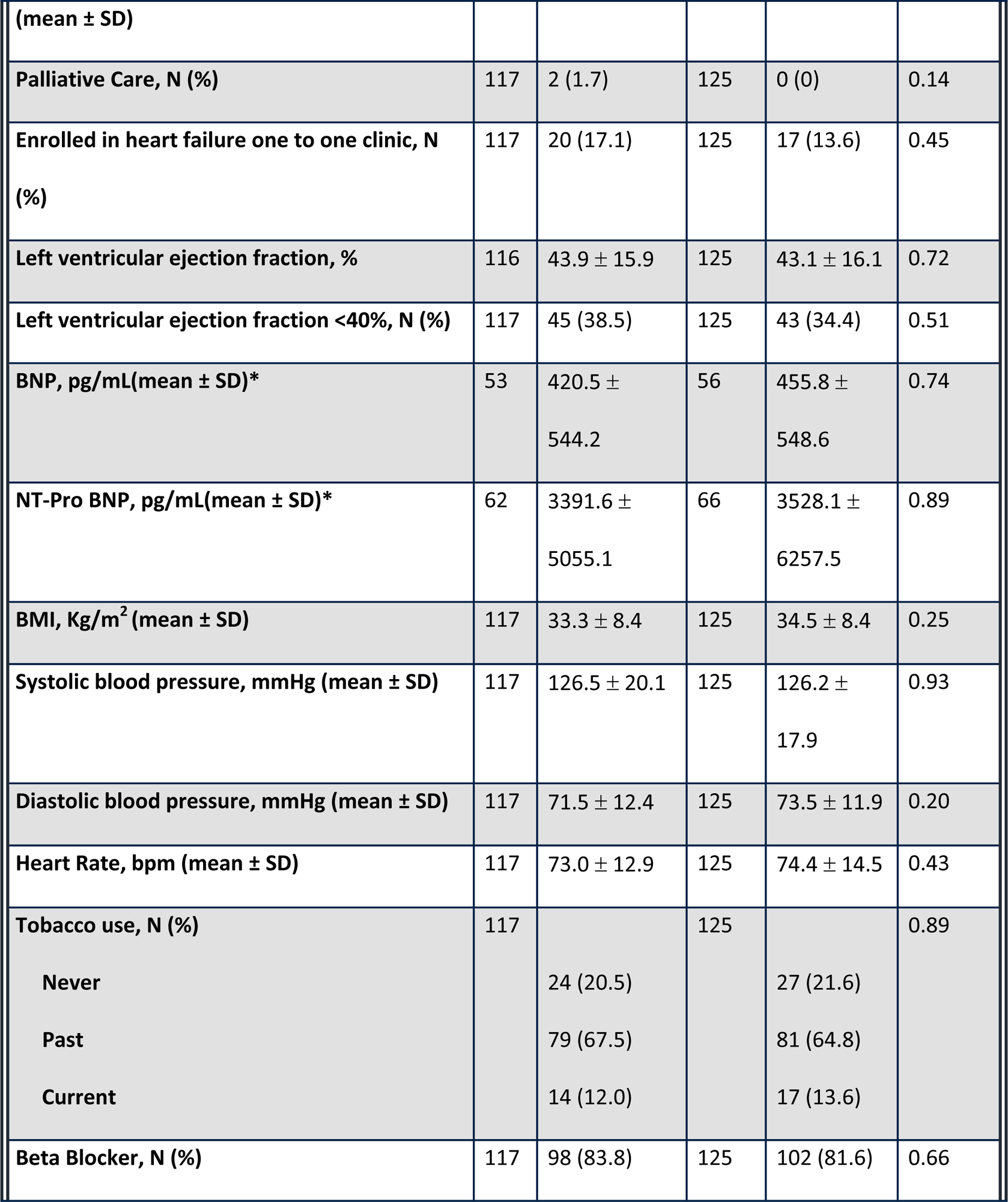
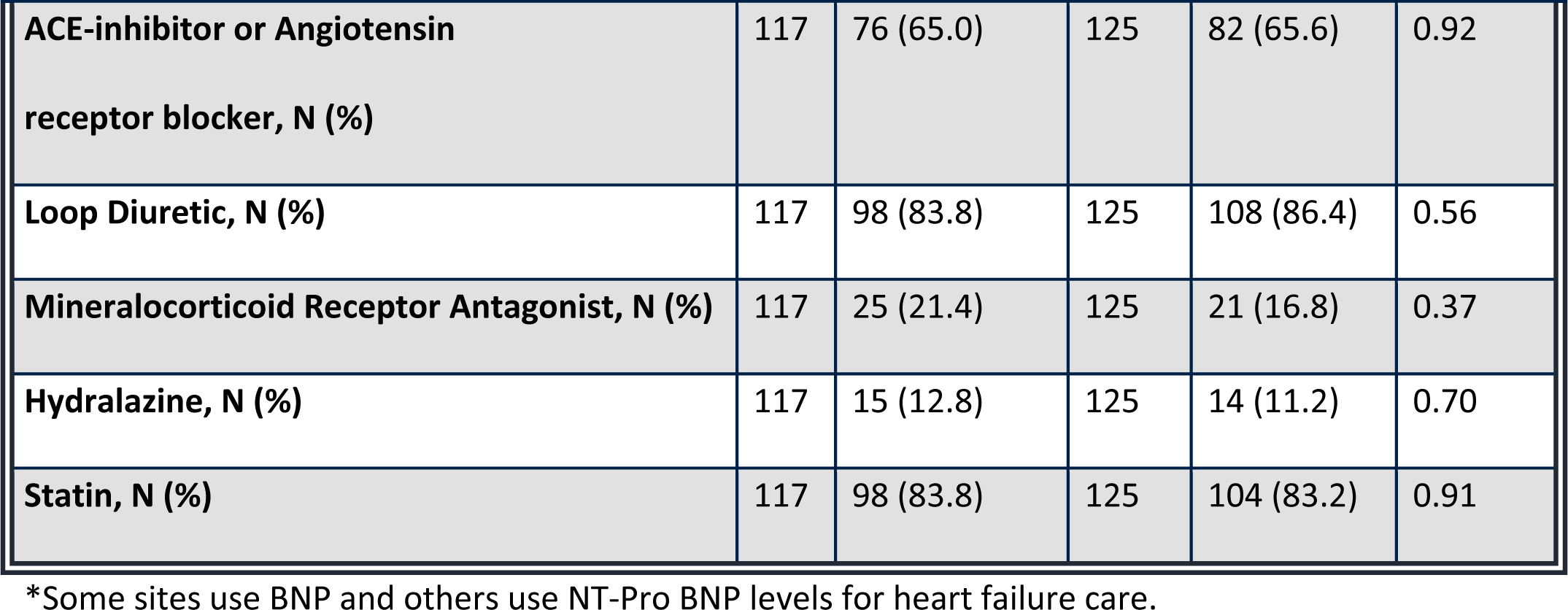
Baseline demographics and clinical characteristics for HF-SMA intervention and usual care arms.

### Primary Outcomes

After 180 days of follow-up, participants randomized to the HF-SMA arm had a greater and significant improvement in the EQ5D-VAS score as compared to those participants randomized to usual care (8.6 ± 16.9 points for HF-SMA versus -0.5 ± 20.7 points for usual care, adjusted p<0.001 for between group comparison) (Figure 2). Participants in the usual care arm did not have a significant improvement in their EQ5D-VAS scores.

**Figure 2:**
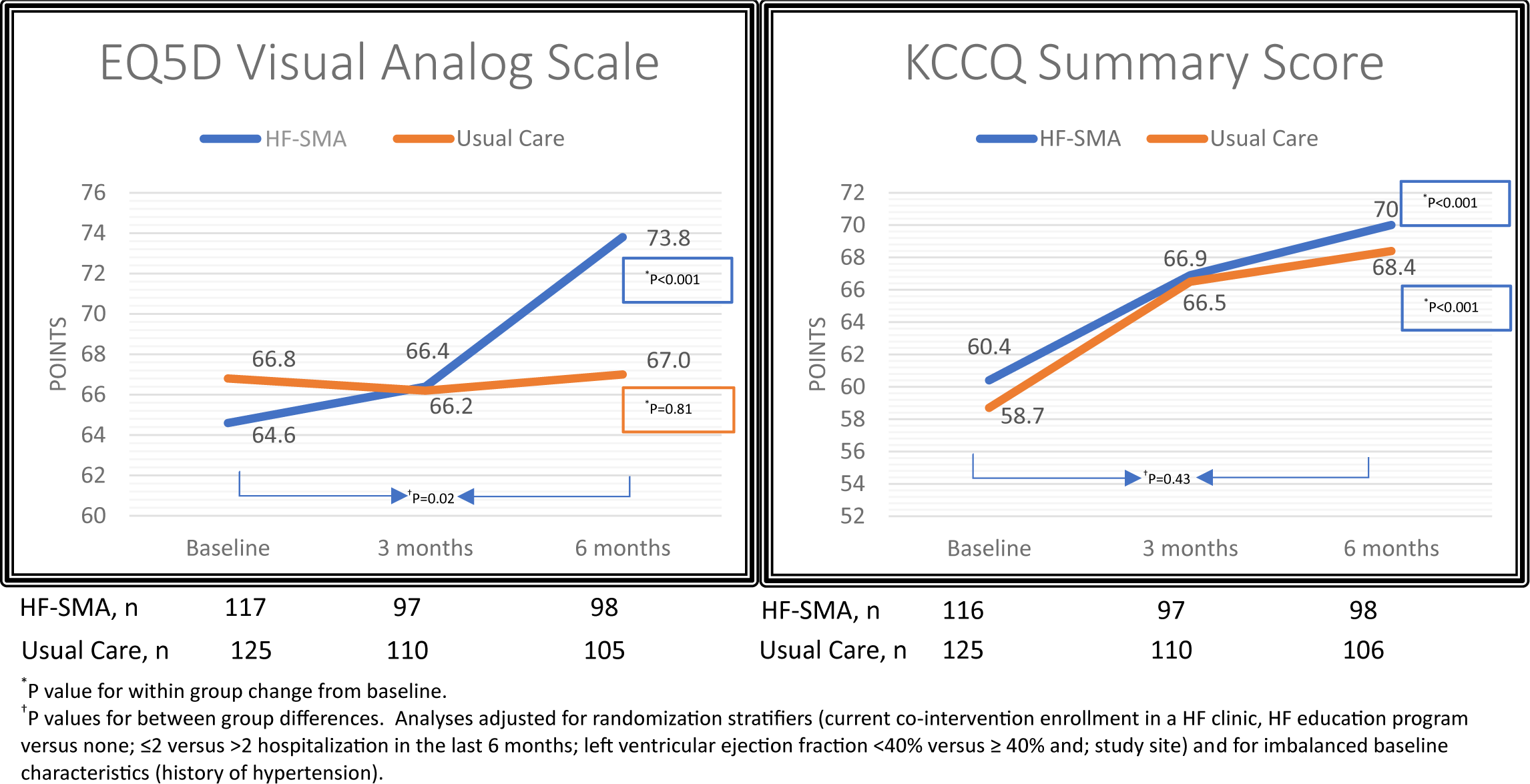
Change in EQ5D Visual Analog Scale and KCCQ Summary Scores over 180 days of follow-up

Conversely, the improvement from baseline in KCCQ Summary Scores were similar (adjusted p=0.43) and significant for both the HF-SMA (8.3 ± 22.9, p<0.001) and the usual care arms (8.5 ± 17.6 points, p<0.001) (Figure 2).

No significant interactions were found between randomization allocation and stratification variables on the change in EQ5D-VAS and KCCQ scores.

### Secondary Outcomes

Of the 3 domains in the Self-Care of Heart Failure Index (SCHFI) scores, participants in the HF-SMA arm had greater and significant improvements in the HF self-care management (HF-SMA 9.4 ± 23.8 points versus usual care 5.2 ± 25.0 points, adjusted p=0.001) and confidence (7.5 ± 22.9 points for HF-SMA versus 2.9 ± 23.2 points for usual care, adjusted p=0.007) domain scores compared to the usual care arm. Patients randomized to the usual HF care arm did not experience a significant improvement in their Confidence scores (p=0.20).

The change in the self-care maintenance domain scores improved significantly from baseline for both the HF-SMA 9.8 ± 17.8 points (p<0.001) and usual care 5.4 ± 16.3 points (p=0.001) arms. The change in self-care maintenance scores did not significantly differ (adjusted p=0.37) between the HF-SMA and usual care arms. (Figure 3)

**Figure 3:**
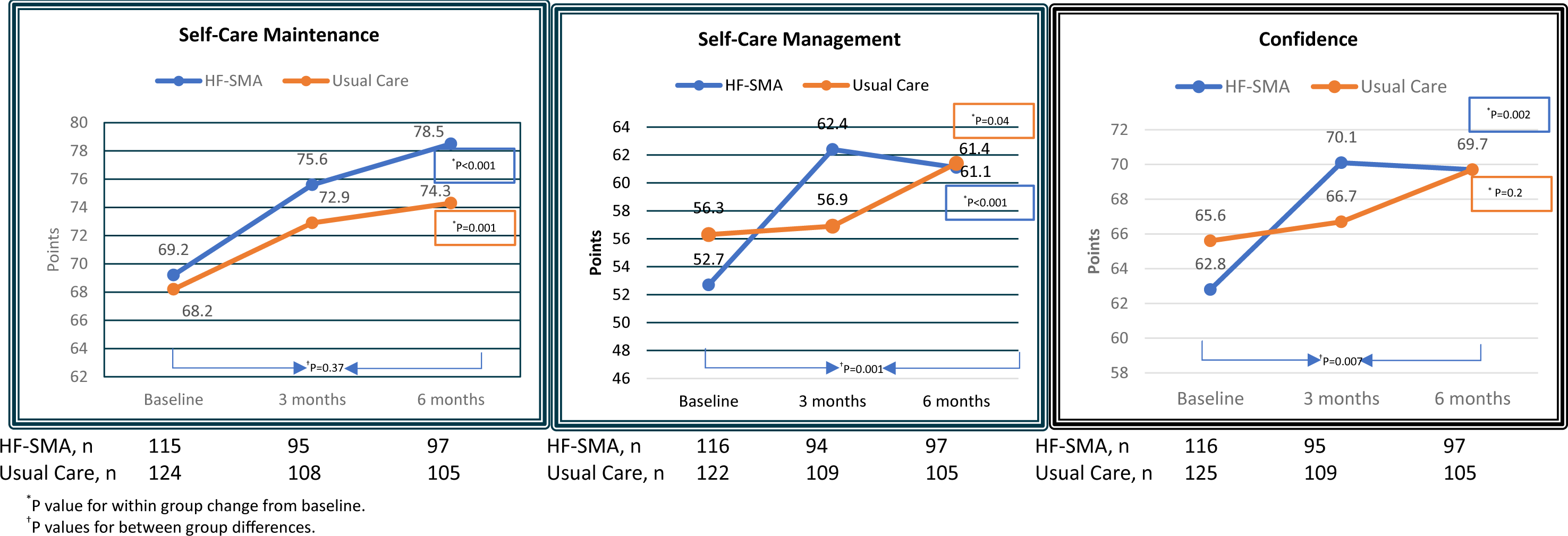
Change in the SCHIFI Scales: Self-care Management, Self-Care Maintenance and Confidence Scales over 180 days of Follow-up

### Rehospitalization or Death

A total of 8 (6.8%) participants in the HF-SMA intervention arm and 9 (7.2%) in the control arm (p=0.91) died during the 180 days of follow-up. A total of 49.6% in HF-SMA and 50.4% in usual HF care were re-hospitalized or died. The adjusted hazard ration (HR) for time to death or hospitalization over the 180 days of follow-up between HF-SMA versus usual HF care was 0.97 ([95% CI 0.68-1.4], p=0.88) (Figure 4).

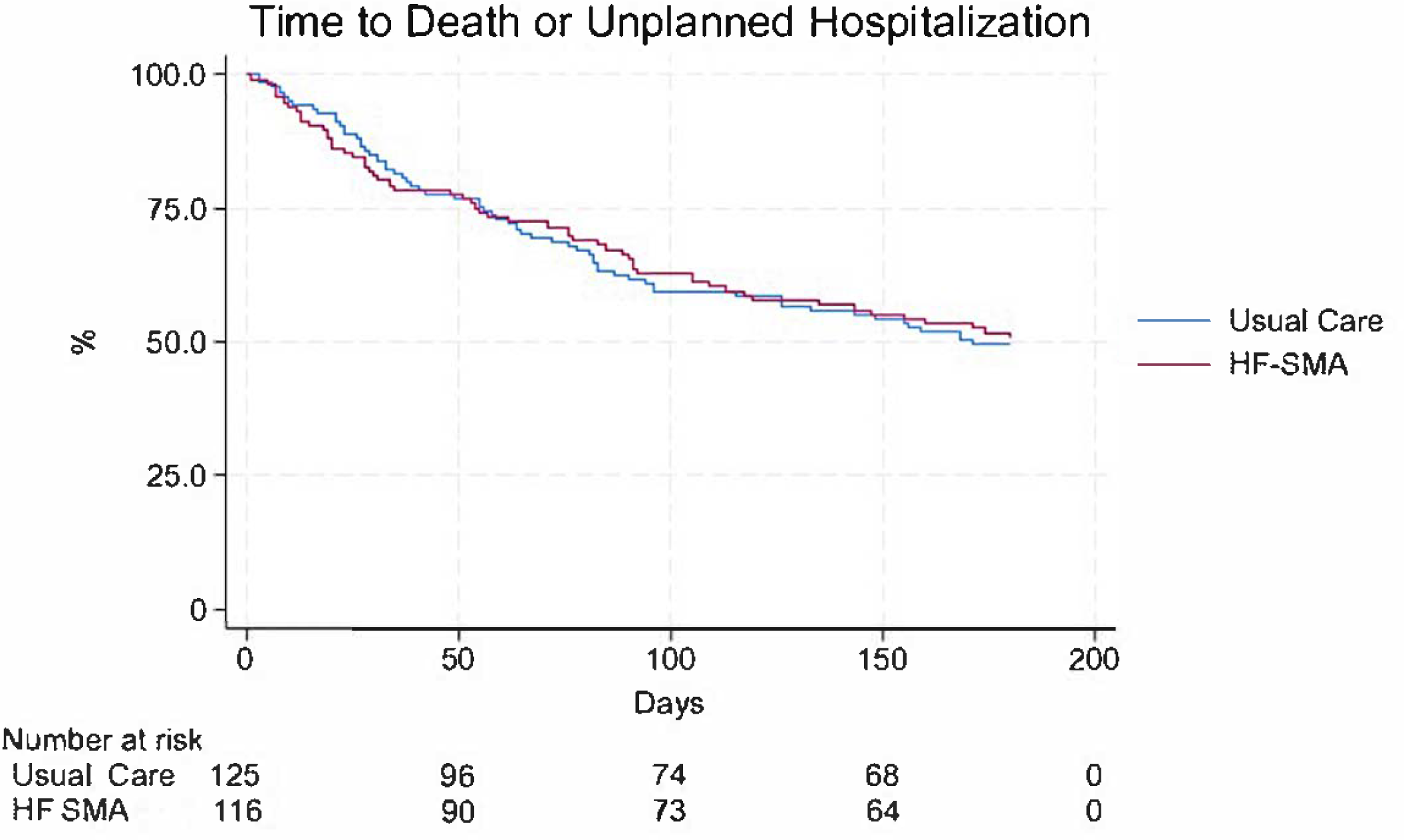

### Co-Interventions

Over the 180 days of follow-up, 44 (37.6%) of the HF-SMA and 50 (40%) of usual care received at least 1 visit in the individual provider HF clinic (mean number of days from enrollment into the HF clinic was 79.5 ± 44.6 days for the HF-SMA arm versus 75.8 ± 47.7 days for usual care, p=0.70).

### Intervention Adherence

Participants randomized to the HF-SMA arm attended an average of 2.5 ± 1.2 out of 4 total HF-SMA visits during the study.

## Discussion

In this RCT of HF-SMA versus usual HF care for patients recently discharged with HF, HF-SMA participants experienced significant and greater improvements in overall health status, and similar improvements in cardiac specific health status compared to participants in the usual HF care over 180-days of follow-up. Process outcomes showed that HF-SMA participants experienced greater improvements in the HF self-care maintenance and confidence subscale scores and similar improvements in health care management scores from baseline compared to usual HF care participants.

Heart Failure is a chronic complex illness that requires a multi-disciplinary care team. Addressing HF in the context of multi-comorbidity necessitates a multidisciplinary approach as treatment plans often require multi-factorial interventions comprised of medication optimization, education on lifestyle modification, and care coordination among various healthcare specialists. The multidisciplinary team structure of the HF-SMA setting permits addressing both HF related and non-HF related factors and shifts the burden of care coordination and multiple care visits from the patient to the healthcare system. The HF-SMA facilitates access to multi-discipline providers in one setting and ensures coordinated communication across disciplines. It facilitates improvement of non-HF symptoms such as psychological well-being, usual activities of daily living, and general discomfort. This may explain the greater and significant improvement in the more generic EQ5D-VAS health status scores for HF-SMA participants versus usual HF care since EQ5D-VAS provides an overall health status that is reflective of the perceived health across multiple health conditions. Of note, participants in the usual HF care arm did not experience a significant improvement in the EQ5D-VAS overall health status scores, while patients from both arms experienced improvements in the cardiac-specific KCCQ health status scores that focus on symptoms and quality of life related to heart failure.

Multidisciplinary interventions play a crucial role in enhancing self-care behavior and confidence through a collaborative holistic approach in which professionals from different disciplines contribute their expertise to educate individuals on healthy lifestyle choices, medication management, and behavioral support to overcome health care system and psychological barriers. However, comprehensive multidisciplinary holistic care often requires the coordination of several care visits with various health care personnel, along with additional time, and travel burden in the traditional individual provider clinical care system. The structure of the HF-SMA program integrates and streamlines in one setting where a multidisciplinary team can provide care and minimizes the coordination burden for both the patient and healthcare team members. The findings that HF-SMA participants achieved a greater improvement in the HF self-care management and confidence scale scores compared to usual care support the notion that self-management education and training were provided better in a HF-SMA setting. Heart Failure self-care scores have been shown to be a valid indicator of risk for admission and death in people with HF.^18^

### Strengths and Limitations

To our knowledge, our RCT represents the first multi-site study to rigorously assess the outcomes of a multidisciplinary team intervention in a group medical clinic setting to improve patient-centered self-reported outcomes of high-risk patients with a recent HF hospitalization. Although literature supports the effectiveness and reduction of health care expenditures with the use of SMAs in other chronic disease states such as diabetes^19–22^, there is limited evidence to support the efficacy of SMAs in severely ill patients with HF. Prior pilot studies evaluating HF-SMA have been retrospective^23^, without active comparators^24^, convenience sample studies,^25^ or a single-site education-based intervention led by a physician and nurse care team;^26^ none of which critically studied a group clinic setting where clinical assessments were made and medications were initiated or adjusted, in addition to self-management training and support.

This trial has limitations. First, we only enrolled 242 of our proposed sample size of 250 participants. However, we met the proposed sample size needed to detect a statistical and clinically relevant difference in our primary outcomes accounting for attrition. Second, only 20% of those who were screened for eligibility and contacted for enrollment agreed to participate in the trial. This may limit the generalizability of our findings, since our sample may represent a more motivated and adherent population that could accept care in a group setting. Third, the duration of follow-up was limited to 6 months; although the trajectory of effects on the EQ5D-VAS and KCCQ summary scores, indicated greater persistent improvements over time for HF-SMA participants versus usual care, the durability of the observed effects beyond 6 months cannot be ascertained. Fourth, our trial was designed primarily to evaluate change in health status and was not adequately powered to evaluate clinical events such as hospitalizations or deaths. Fifth, participants were mostly older white men, which is representative of the Veteran HF population, but may limit the generalizability to women.

### Conclusion

Both HF-SMA and usual care in HF participants achieved significant improvements in cardiac health status, but only a team approach through HF-SMA achieved significant improvements in overall health status. A larger population and a longer follow-up time are needed in future studies to evaluate re-hospitalization and death outcomes.

## Other information

### Registration

ClinicalTrials.gov ID: NCT02481921

### Disclaimer

The views expressed are those of the authors and do not represent the views of the Department of Veterans Affairs

### Disclosure

None

## Data Availability

The data that support the findings of this study are available from the corresponding author upon reasonable request.

